# Meta-analysis of rapid direct-to-PCR assays for the qualitative detection of SARS-CoV-2

**DOI:** 10.1101/2021.05.07.21256745

**Authors:** T.A. Reginald, S. Grippon, M.J. Coldwell, H. Chen, L. Koh, U. Jan, A. Sanchez-Bretano, D. Borley, P. Oladimeji, N. Moore, S.P. Kidd, J.E. Martin

## Abstract

Infection with severe acute respiratory syndrome coronavirus 2 (SARS-CoV-2) and the ensuing COVID-19 pandemic present significant challenges to current diagnostic and therapeutic patient care pathways including whether new *in vitro* diagnostic tests can accurately identify and rule out current SARS-CoV-2 infection.

The gold standard diagnostic test to identify a current SARS-CoV-2 infection is a central laboratory-based molecular assay employing reverse transcription polymerase chain reaction (RT-PCR) with very high accuracy of detection; however, which typically requires 1-2 days turn-around for results. Rapid RT-PCR assays and systems have been developed which can be deployed locally (near-patient or point of care (POC)), provide faster results and not impact on already stressed central laboratory capacity. Rapid test results can be returned within the same clinical encounter, facilitating timely decisions that optimise the patient care pathway and support more rapid COVID-19 diagnosis, isolation and contract tracing activities^1^.

Direct-to-PCR is an evolution of RT-PCR in which the patient sample is added directly to an amplification reaction without being subjected to prior nucleic acid extraction, purification, or quantification to reduce the time and monetary resources required to process samples. Rapid, direct-to-PCR systems further increase the speed of testing by combining rapid PCR instruments with direct-to-PCR assays, to generate results in less than two hours.

This appears to be the first meta-analysis assessing the accuracy of rapid direct-to-PCR in the detection of SARS-CoV-2. In total, 1,144 unique records were identified and screened using search string evaluation, 49 full-text reports and/or supplemental materials were assessed for inclusion. This resulted in 16 studies, reporting 22 datasets with 5322 patient samples (of which 2220 were identified as positive according to centralised laboratory testing) included in the analysis.

The overall percentage agreement (OPA) between the rapid direct RT-PCR and gold standard centralised laboratory RT-PCR was 95.10% with 91.22% positive percent agreement (PPA) and 98.16% negative percent agreement (NPA). When compared to commercially available tests were considered, these were assessed to be 96.95% OPA, 94.78 % PPA and 98.34 % NPA. Furthermore, the Cohens kappa statistical coefficient k = 0.94 (0.96 for commercial only), and Youden Index = 0.893 (0.924 for commercial only) indicate an almost perfect agreement. These results therefore indicate that direct-to-PCR assays performance is equivalent to the standard centralised laboratory PCR systems for the detection of SARS-CoV-2.

**Objectives:** To assess the efficacy of rapid direct-to-PCR assays and systems for the detection of SARS-CoV-2 in the hospital, care home and medical research population from November 2020 to July 2021.

**Search methods:** Initial electronic searches of the Cochrane COVID-19 Study Register (which includes daily updates from PubMed and Embase and preprints from medRxiv and bioRxiv) were undertaken on the 30^th^ of April 2021, with a further search undertaken on 8^th^ July 2021 (PRISMA flow diagram, Figure 2).

**Selection criteria:** Studies, published in English, of subjects with either suspected SARS-CoV-2 infection, known SARS-CoV-2 infection or known absence of infection, or those who were being screened for infection were included. Commercially available and research use only rapid direct-to-PCR assays (without RNA extraction and purification reporting results within two hours) were included in the study.

**Data collection, extraction and analysis:** Studies were screened independently, in duplicate with any disagreements resolved by discussion with a third author. Study characteristics were extracted by one author and checked by a second; extraction of study results and assessments of risk of bias and applicability were undertaken independently in duplicate.

Where studies were not publicly available, sites that undertook in-service evaluations of rapid direct-to-PCR system were contacted and asked to supply anonymised datasets. Both reviewers independently performed data extraction and verification and calculated 2×2 contingency tables with the number of true positives, false positives, false negatives and true negatives. They resolved any disagreements by discussion and by review with the third reviewer.

**Main results:** In total, 22 study cohorts were included (described in 16 study reports, including 5 unpublished reports), reporting results for 5322 samples (of which 2220 were confirmed SARS-CoV-2, as determined by central laboratory testing). Studies were mainly from Europe and North America and evaluated eight commercially available direct-to-PCR assay kits/cartridges, and six developed from other reagents.

**Conclusions:** This appears to be the first meta-analysis assessing the accuracy of rapid direct-to-PCR in the detection of SARS-CoV-2. In total, 1,144 unique records were identified and screened using search string evaluation, 49 full-text reports and/or supplemental materials were assessed for inclusion. This resulted in 16 studies reporting 21 datasets with 5322 patient samples (2220 positive) included in the analysis.

The overall agreement between the commercially available rapid direct RT-PCR and gold standard centralised laboratory RT-PCR was 96.9% with 94.8% PPA and 98.4% NPA. Furthermore, the Cohe_n_s kappa statistical coefficient k = 0.96, indicating an almost perfect agreement and Youden Index = 0.93. These results show that direct-to-PCR assays performance is equivalent to the gold standard centralised laboratory RT-PCR systems for the detection of SARS-CoV-2.

**Plain language summary:** *What is a rapid direct-to-PCR test for diagnosing COVID-19?:* Rapid direct-to-PCR tests are rapid tests that aim to confirm or rule out the presence of SARS-CoV-2 within 2 hours without complicated processing of the sample.

*How accurate is a rapid direct-to-PCR test for diagnosing COVID-19?:* We compared the accuracy of rapid direct-to-PCR tests with gold standard centralised laboratory RT-PCR for the detection of SARS-CoV-2 and found that direct-to-PCR was as accurate as standard RT-PCR assays.

*Why is this question important?:* People with suspected COVID-19 need to know quickly whether they are infected, so that they can self-isolate, inform close contacts and possibly receive treatment. Currently, COVID-19 infection is confirmed by a laboratory test called RT-PCR, which uses specialist equipment and often takes at least 24 hours to produce a result. If they are accurate, faster diagnosis could allow people to take appropriate action more rapidly, with the potential to reduce the spread of COVID-19.^1^

*What did we aim to find out?:* Our goal was to determine if commercially available and research use rapid direct-to-PCR tests are accurate enough to detect SARS-CoV-2 in comparison to gold standard laboratory RT-PCR.

*What did we do?:* We looked for studies that measured the accuracy of any commercially produced and research use rapid direct-to-PCR tests, in people tested for COVID-19 using RT-PCR. People could be tested in hospital or in the community. Studies could test people with or without symptoms. Tests had to use minimal equipment, be performed safely without risking infection from the sample, and have results available within two hours of the sample being collected.

*What we found?:* We include 22 studies in the review. They investigated a total of 5322 nose or throat samples; COVID-19 was confirmed in 2220 of these samples. The studies investigated 15 different direct-to-PCR tests. They took place mainly in Europe and North America.

*What did we find?:* Although overall results for diagnosing and ruling out COVID-19 were good (91.2% of infections correctly diagnosed and 98.3% correctly ruled out), we noted a difference in COVID-19 detection between tests, especially those available as commercial kits versus ones assembled from reagents from different sources. However, we cannot be certain about whether results will remain the same in a real-world setting. We could not investigate differences in people with or without symptoms, nor time since symptoms-onset because the studies did not consistently provide enough clinical information about their participants.

*How reliable were the results of the studies?:* In general, the studies included followed rigorous methods, in accordance with the tests intended use to detect COVID-19 and included at least two independent results to confirm or rule out COVID-19 infection. The results from different test brands varied and few studies compared multiple rapid-PCR tests. Most of the studies did not provide sufficient information to determine whether the detection levels would vary in people with COVID-19 symptoms versus without symptoms.

*What does this mean?:* On average the rapid direct-to-PCR were shown to be equivalent to gold standard laboratory-based RT-PCR tests and several direct-to-PCR tests show very high accuracy. However, for most of the tests, more evidence is needed particularly in people without symptoms, on the accuracy of repeated testing, and testing in non-healthcare settings such as schools (including self-testing).

## Background

Infection with severe acute respiratory syndrome coronavirus 2 (SARS-CoV-2) and the ensuing COVID-19 pandemic present significant diagnostic and therapeutic challenges. These range from: understanding the value of signs and symptoms in predicting possible infection; assessing whether existing biochemical and imaging tests can identify infection or people needing critical care; and evaluating whether *in vitro* diagnostic tests can accurately identify and rule out current SARS-CoV-2 infection, and identify those with past infection, with or without immunity.^1^

The standard diagnostic test to identify a SARS-CoV-2 infection is a central laboratory-based molecular assay using reverse transcription polymerase chain reaction (RT-PCR).

Direct-to-PCR is an evolution of PCR in which a sample is added directly to an amplification reaction without being subjected to prior nucleic acid extraction, purification, or quantification. It allows for maximum source quantities of RNA to be targeted, minimises opportunities for error and contamination, and may be less expensive, provide results more quickly and not require the same laboratory capacity, avoiding the need for centralised testing facilities.

Rapid direct-to-PCR systems further increase the speed of testing by combining rapid-PCR instruments with faster cycling times and data analysis with direct-to-PCR assays to generate results in less than 2 hours.

If sufficiently accurate, rapid tests returned within the same clinical encounter can facilitate timely decisions concerning the need for isolation and contract tracing activities^1^ and to facilitate novel patient care pathways for the rapid diagnosis and treatment of COVID-19 and supporting infection prevention and control.

### Description of the condition

COVID-19 is the disease caused by infection with the SARS-CoV-2 virus. The target condition for this meta-analysis is current SARS-CoV-2 infection diagnosed by a positive RT-PCR detection of the SARS-CoV-2 RNA.

### Description of the test

Direct-to-PCR is an evolution of RT-PCR in which a sample is added directly to an amplification reaction without being subjected to prior nucleic acid extraction, purification, or quantification. It allows for maximum quantities of source RNA to be targeted, minimises opportunities for error and contamination, and reduces the time and monetary resources required to process samples. Rapid direct-to-PCR systems further increase the speed of testing by combining rapid-PCR instruments with faster cycling times and data analysis with direct-to-PCR assays, generating results in less than 2 hours.

### Index test(s)

The primary consideration for the eligibility of tests for inclusion in this review is that they were aimed at the detection of SARS-CoV-2 infection in samples without being subjected to prior nucleic acid extraction, purification, or quantification and with results in less than 2 hours.

### Clinical pathway

Patients may be tested for SARS-CoV-2 when they present with symptoms, have had known exposure to a confirmed case, or are part of a screening program, with no definite known exposure to SARS-CoV-2. The standard approach to diagnosis of SARS-CoV-2 infection is through laboratory-based testing of swab samples taken from the upper respiratory (e.g., nasopharynx, oropharynx) or lower respiratory tract (e.g., bronchoalveolar lavage or sputum) with RT-PCR.^1^

### Rationale

It is essential to understand the clinical accuracy of tests and clinical features to identify the best way they can be used in different settings to develop effective diagnostic and management pathways for SARS-CoV-2 infection and disease. Estimates of accuracy from these reviews will help inform diagnosis, screening, isolation, and patient-management decisions.^1^

## Objectives

To assess the efficacy of rapid direct-to-PCR systems for the qualitative diagnosis of SARS-CoV-2 in the hospital, care home and medical research population from November 2020 to July 2021.

## Methods

### Criteria for considering studies for this review

#### Methods

MeSH terms used in the search string:

1. Covid-19[MeSH Terms] OR SARS-CoV-2[MeSH Terms]
2. “direct to pcr” OR direct OR rapid OR “extraction free” OR “extraction-free”
3. “RT-qPCR” OR “qRT-PCR” OR “RT-PCR” OR “-PCR” OR “real-time PCR”
4. #2 AND #3
5. #1 AND #4

The review was conducted following the Cochrane Rapid Review process and workflow (v.4) and we have referenced and followed the approach of the 2021 Cochrane Rapid, point-of-care antigen and molecular-based tests for diagnosis of SARS-CoV-2 infection review^1^. We conducted the preliminary literature search for existing or ongoing systematic reviews using the Cochrane Library and estimated review feasibility. The protocol was drafted, and a team of systematic review authors screened the titles and abstracts of all records retrieved from the literature searches following the search string optimisation described in the appendix. Two review authors independently screened studies and a third, senior review author resolved any disagreements. We tagged all records selected and obtained the full texts for all studies flagged as potentially eligible. Two review authors independently screened the full texts, and we resolved any disagreements on study inclusion through discussion with a third review author.

#### Assessment of Risk of Bias

The authors assessed and discussed the risk of bias and applicability concerns using the RoB 2.0 algorithm tailored to this review^2^, and concluded that the categorization ‘some concerns’ was appropriate (Figure 1).

**Figure 1.**
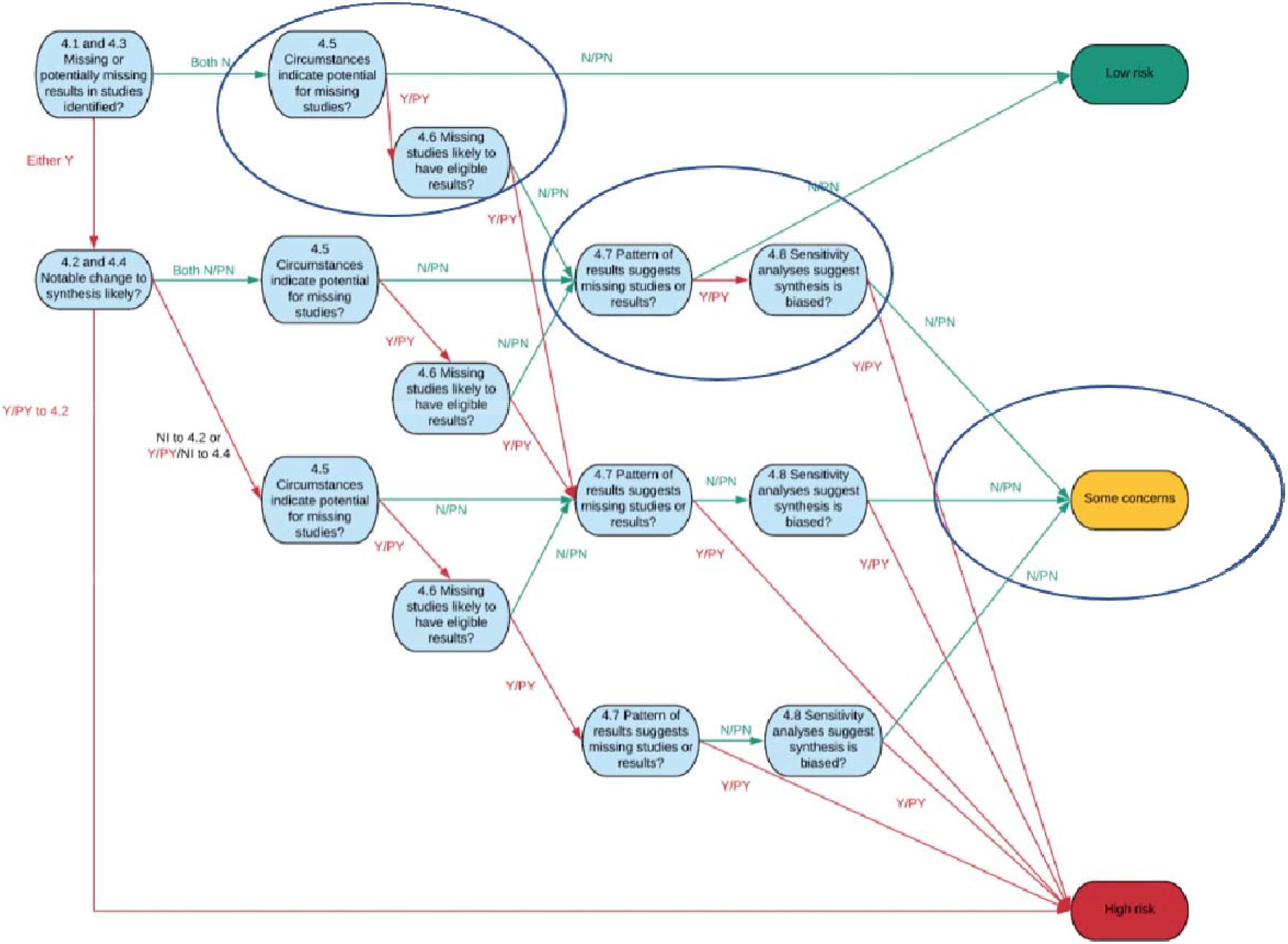
A tool for assessing Risk of Bias due to Missing Evidence in a synthesis (ROB-ME)

**Figure 2.**
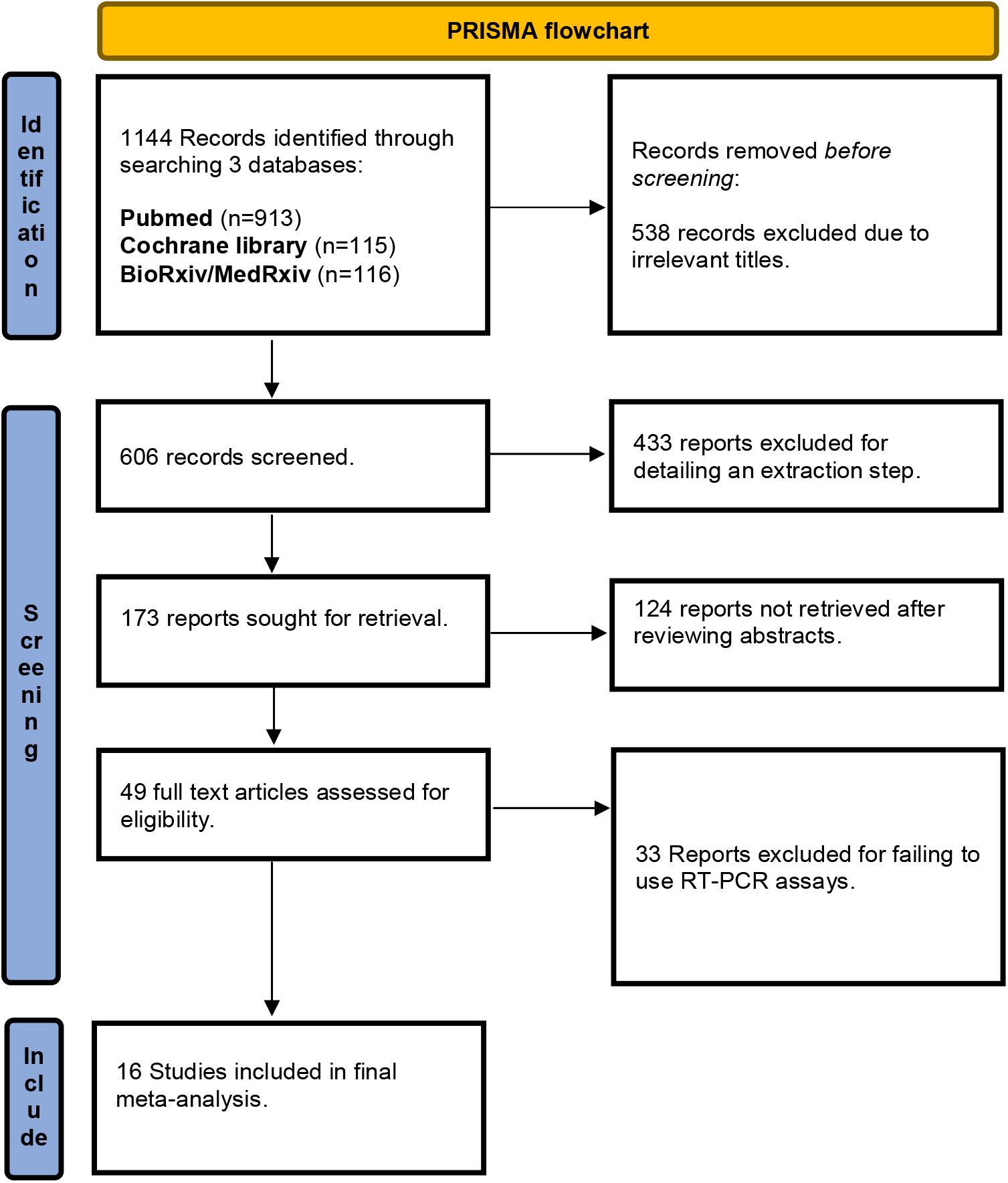
PRISMA flowchart.

#### Statistical analysis and data synthesis

We analysed rapid direct-to-PCR assays and computed estimates by summing the counts of true positives, false positives, false negatives, and true negatives across 2×2 contingency tables and verifying the supplemental information in publications or source data for the unpublished studies. We made comparisons between analyses using the rapid direct-to-PCR assay and standard centralized lab assay. Any discrepant results were adjudicated using a third reference assay.

#### Assessment of reporting bias

We have indicated where we were aware that study results were available but unpublished and have created a Funnel Plot which enables studies with the potential to bias the overall meta-analysis to be identified (Figures 4 and 5).

#### Duration

The minimum study duration was one month, and the target population encompassed symptomatic and asymptomatic residents and staff in care homes, patients in hospitals, healthcare workers and medical researchers.

#### Intervention

In an ideal situation, paired nasopharyngeal, oropharyngeal or nasal patient swabs would be collected. One swab from each pair then analysed using the rapid direct-to-PCR system and the other swab tested using the gold standard centralised laboratory PCR system. If the results were discordant, a third reference PCR system would be utilised to provide the definitive diagnosis. However, in several studies, a single sample was split for assay in two (sometimes more) assays, not necessarily at the same time, meaning that assays were carried out on samples which may have undergone further freeze-thaw or been processed at vastly different times.

#### Direct to PCR systems assessed

VitaPCR™ NAAT

SureFast^®^ SARS-CoV-2 PLUS Test

DiaSorin Molecular Simplexa COVID-19 Direct EUA

exsig COVID-19 Direct

PROmate COVID-19 (1G and 2G)

Hyris Kit Simplexa

#### Comparator PCR systems

The assay established by Corman et al (2020)^3^

The New York SARS-CoV-2 Real-time Reverse Transcriptase (RT)-PCR Diagnostic EUA Panel (Modified 96 CDC assay)

Abbott ID NOW

Hologic Panther Fusion® SARS-CoV-2 EUA Hologic Panther

GenMark ePlex SARS-CoV-2 EUA panel

Cepheid

Alinity (Abbott)

M2000 (Abbott)

Roche Cobass

Genesig COVID-19

#### Search methods for identification of studies

Electronic databases:

Pubmed and Cochrane COVID-19 study registry from December 1^st^ 2019 to July 8^th^ 2021

#### Other searches

Public Health England Technical Validation Group

Manufacturer websites

Unpublished NHS validation studies

#### Screening

Two reviewers screened all titles and abstracts for eligibility and no non-English titles were reviewed.

### Data collection and analysis

#### Software

Excel and Sharepoint, R (mada package), RevMan 5.0

#### Data extraction

Conducted with a pilot-tested form by two reviewer and verified by a third using Excel to record the following:

Sample number

Exsig assay result

Exsig assay Cq value

Exsig assay IC value

Comparator assay result

Comparator assay Cq value

Comparator IC

Outcomes assessed: Cq values to determine SARS-CoV-2

Numerical data for outcomes of interest: Cq values

#### Contacting study authors

Authors were contacted if contingency table or source data was unavailable from the publication.

#### Data management

Excel and Sharepoint

#### Data synthesis

Published contingency tables were verified from results in the supplemental information and used to calculate overall (OPA), positive (PPA) and negative (NPA) percent agreement for published records. Source data was extracted and verified by two reviewers for unpublished studies to calculate the contingency table and then calculate OPA, PPA and NPA as above.

#### Analysis

OPA, PPA, NPA, Cohen kappa (statistical coefficient to measure the agreement) and the Youden Index were calculated.

## Results

### Description of studies

#### Results of the search

In all, 10,957 unique records (published or preprints) were initially screened for inclusion, with 1,144 identified through the, aforementioned, MeSH terms. Finally, 606 records were selected for further assessment by review of the abstract and/or full-text reports and 49 studies for review of full-text reports and / or supplemental materials. Sixteen reports were selected for requiring assessment for inclusion in this review. See Figure 2 for the PRISMA flow diagram of search and eligibility results.

#### Included studies

Sixteen studies were included which reported 22 cohort datasets including 5322 patient samples of which 2220 were SARS-CoV-2 positive specimens.

### Effects of interventions/results of the synthesis

#### Outcomes

A total of 10,957 unique records were identified and screened using search string evaluation. 49 full-text reports and/or supplemental materials were assessed for inclusion. 16 studies reporting 22 datasets with 5322 patient samples (2220 positive) were included in the analysis. There were 5075 concordant and 247 discordant results as described in the contingency table below (Table 3).

**Table 3.** Summary contingency table.

|  |  | Central |  |
| --- | --- | --- | --- |
|  |  | + | - |
| Direct | + | 2025 | 52 |
|  | - | 195 | 3050 |

The overall agreement between the rapid direct PCR and standard centralised laboratory PCR was 95.36% with 91.22% positive percent agreement and 98.32% negative percent agreement and is summarized in Figure 3.

**Figure 3.**
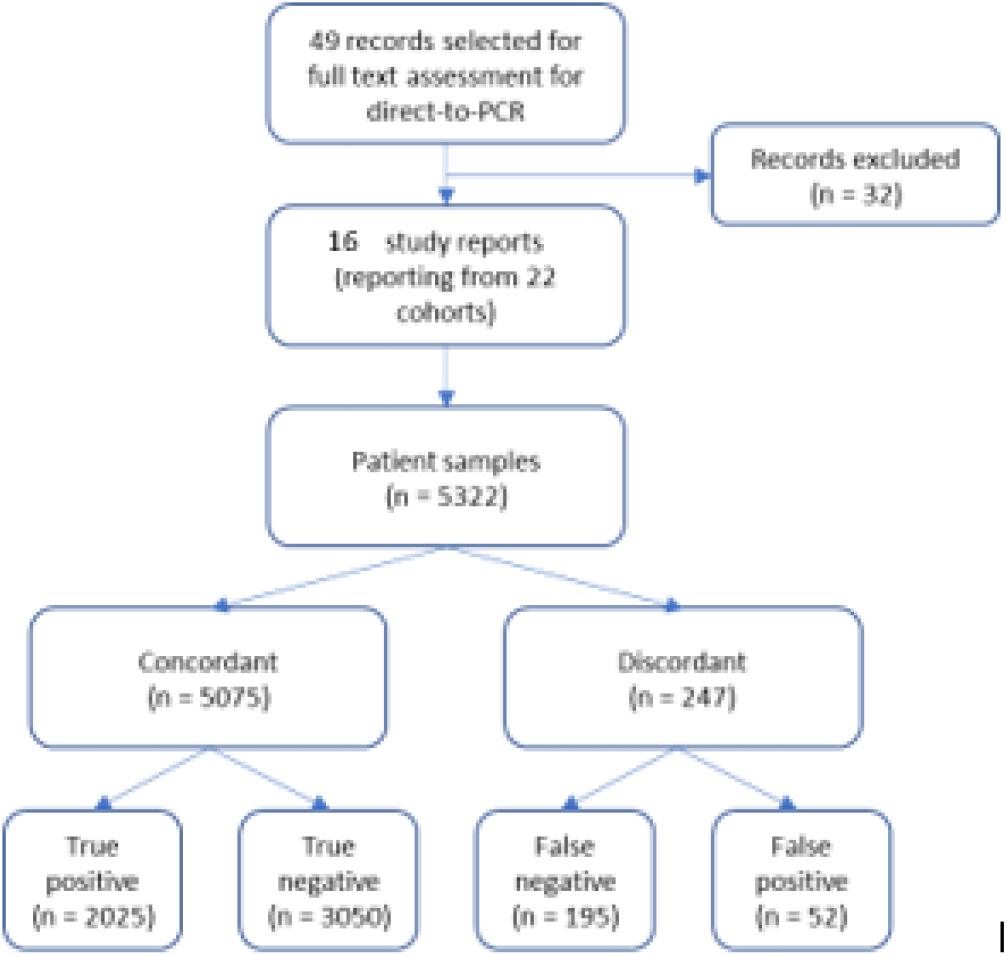
Summary of concordance.

**Figure 4.**
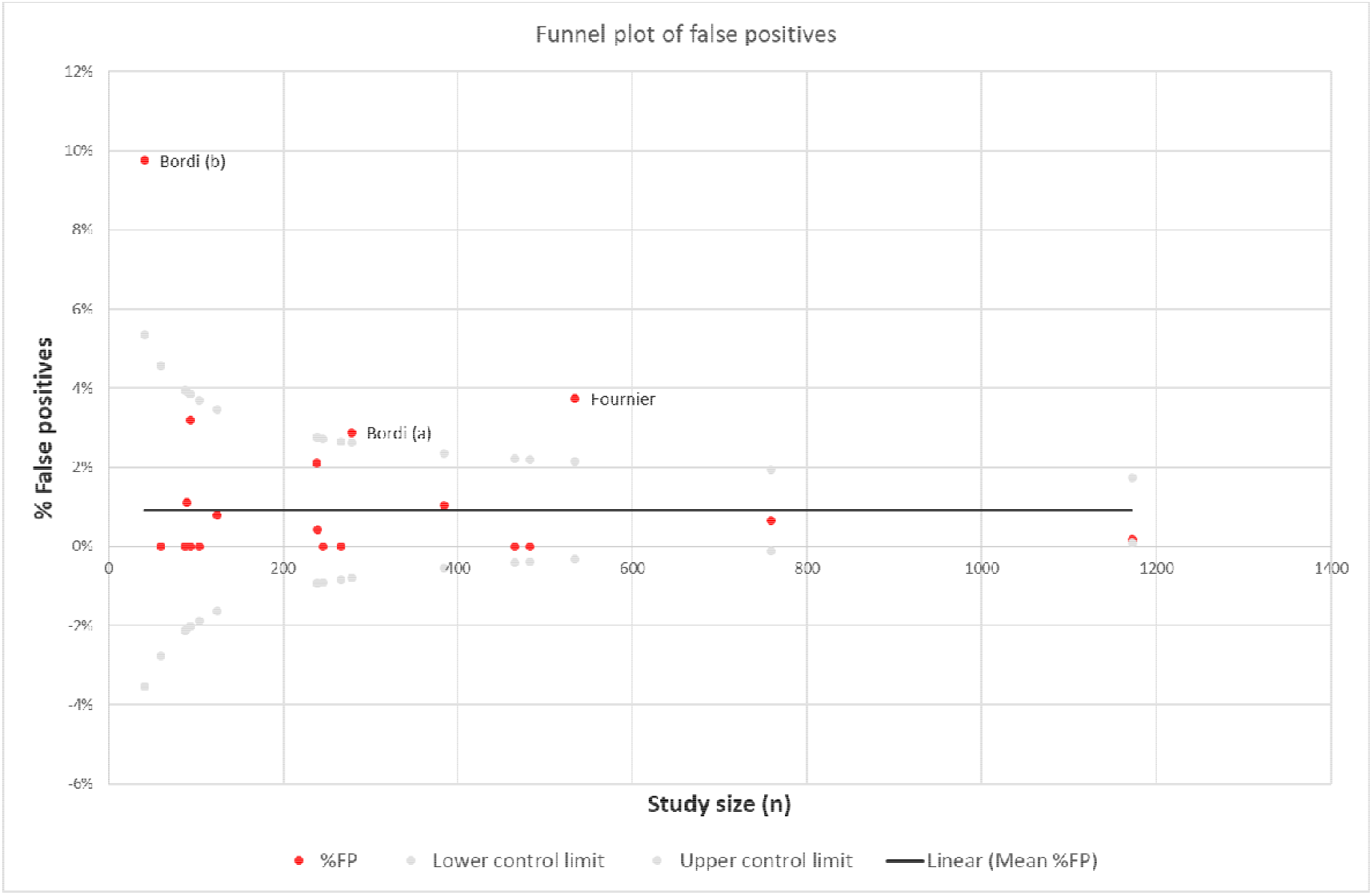
Funnel plot of false positives in studies under analysis. Red dots represent % false positive in the study, and the black line the mean false positive rate from all studies. The lower and upper control limits (grey dots) are plotted at the (mean +/- 3x the standard error) for that study size. Those plotted outside the control limits may be biasing the meta-analysis.

**Figure 5.**
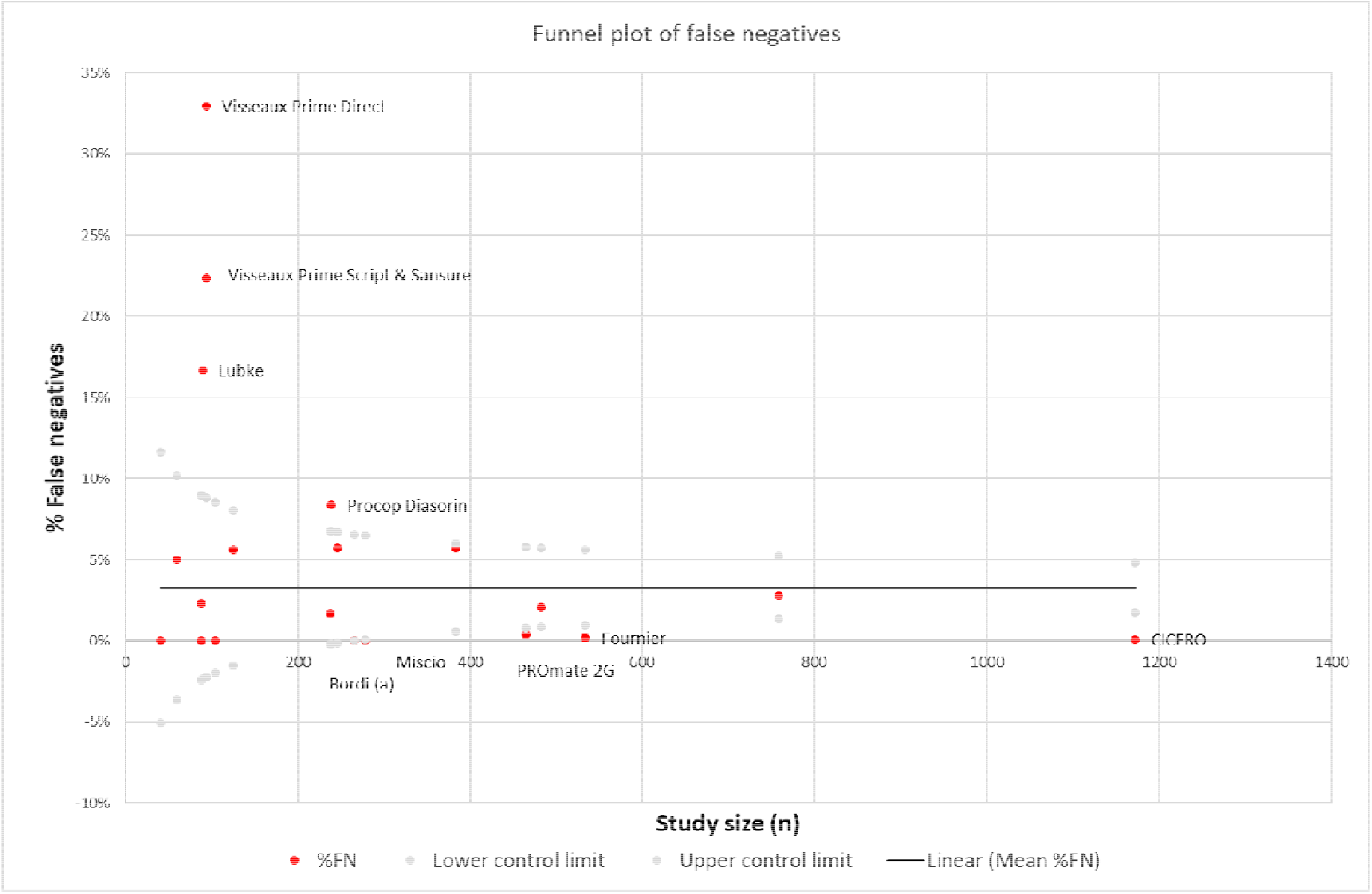
Funnel plot of false negatives in studies under analysis. Red dots represent % false positive in the study, and the black line the mean false negative rate from all studies. The lower and upper control limits (grey dots) are plotted at the (mean +/- 3x the standard error) for that study size. Those plotted outside the control limits may be biasing the meta-analysis

The results and outcomes for each study are shown in Table 4 and an expanded analysis and references provided in the supplemental materials.

**Table 4.** Summary of cohort results.

| Name of study | Samples | True Positive | False Positive | False Negative | True Negative | PPA | NPA | OPA |
| --- | --- | --- | --- | --- | --- | --- | --- | --- |
| Bordi et al (2020a)<sup>4</sup> | 278 | 99 | 8 | 0 | 171 | 100.00% | 95.53% | 97.12% |
| Bordi et al (2020b)<sup>5</sup> | 41 | 21 | 4 | 0 | 16 | 100.00% | 80.00% | 90.24% |
| Byrnes et al<sup>6</sup> | 246 | 82 | 0 | 14 | 150 | 85.42% | 100.00% | 94.31% |
| CICERO<sup>7</sup> | 453 | 154 | 0 | 1 | 298 | 99.35% | 100.00% | 99.78% |
| Clinical evaluation of Exsig Direct<sup>8</sup> | 88 | 43 | 0 | 0 | 45 | 100.00% | 100.00% | 100.00% |
| Clinical evaluation of PROmate 1G<sup>8</sup> | 88 | 43 | 0 | 2 | 43 | 95.56% | 100.00% | 97.73% |
| Clinical evaluation of PROmate 2G<sup>9</sup> | 465 | 181 | 0 | 2 | 282 | 98.91% | 100.00% | 99.57% |
| NHS evaluation of Exsig Direct<sup>10</sup> | 478 | 131 | 0 | 8 | 339 | 94.24% | 100.00% | 98.33% |
| Fournier et al (2020)<sup>11</sup> | 534 | 155 | 20 | 1 | 358 | 99.36% | 94.71% | 96.07% |
| Jorgensen et al (2021)<sup>12</sup> | 60 | 27 | 0 | 3 | 30 | 90.00% | 100.00% | 95.00% |
| Kellner et al (2021)<sup>13</sup> | 384 | 139 | 4 | 22 | 219 | 86.34% | 98.21% | 93.23% |
| Liotti et al (2021)<sup>14</sup> | 125 | 47 | 1 | 7 | 70 | 87.04% | 98.59% | 93.60% |
| Lubke et al (2020)<sup>15</sup> | 90 | 74 | 1 | 15 | 0 | 83.15% | 0.00% | 82.22% |
| Miscio et al (2020)<sup>16</sup> | 266 | 63 | 0 | 0 | 203 | 100.00% | 100.00% | 100.00% |
| Procop et al (2021) - Cepheid<sup>17</sup> | 238 | 163 | 5 | 4 | 66 | 97.60% | 92.96% | 96.22% |
| Procop et al (2021) - DiaSorin<sup>17</sup> | 239 | 148 | 1 | 20 | 70 | 88.10% | 98.59% | 91.21% |
| TVG evaluation of PROmate 1G<sup>18</sup> | 759 | 221 | 5 | 21 | 512 | 91.32% | 99.03% | 96.57% |
| Visseaux et al (2021)- Prime direct<sup>19</sup> | 94 | 38 | 3 | 31 | 22 | 55.07% | 88.00% | 63.83% |
| Visseaux et al (2021) - Prime script<sup>19</sup> | 94 | 48 | 0 | 21 | 25 | 69.57% | 100.00% | 77.66% |
| Visseaux et al (2021) - Sansure<sup>19</sup> | 94 | 48 | 0 | 21 | 25 | 69.57% | 100.00% | 77.66% |
| Zhen et al (2020) - DiaSorin<sup>20</sup> | 104 | 51 | 0 | 0 | 53 | 100.00% | 100.00% | 100.00% |
| Zhen et al (2020) - Genmark<sup>20</sup> | 104 | 49 | 0 | 2 | 53 | 96.08% | 100.00% | 98.08% |
| Totals | 5322 | 2025 | 52 | 195 | 3050 | 91.22% | 98.32% | 95.36% |

Abbreviations: PPA – positive percentage agreement; NPA – negative percentage agreement; OPA – overall percentage agreement

#### Funnel plots to examine reporting bias

Given the wide range of cohort sizes in the studies under analysis, the false positive and false negative rates were subjected to funnel plot analysis to identify those, where the false positives (Figure 4) or false negatives (Figure 5) may be contributing to bias.

As can be observed, several studies fall outside the control limits on either false positive or false negative rates, and in one instance both. This warrants further investigation and deeper analysis of comparator assays and whether a resolver assay or other means should have been used to aid in assigning discordant results. Furthermore, what attempts have the authors made to examine those samples where the Cq value is high and therefore indicative of an assay working at the very limits of detection?

On false positives, then we are able to examine the study by Fournier et al (2020)^11^ in more detail as the symptoms from these 20 patients are reported. These patients are giving a positive result with the VitaPCR assay, which detects the N gene, but negative with an E gene-specific kit. Two asymptomatic patients were tested a few days later and found to be negative, but of the other 18, they exhibited at least one of the hallmark symptoms of Covid-19, as summarised in Figure 6.

**Figure 6.**
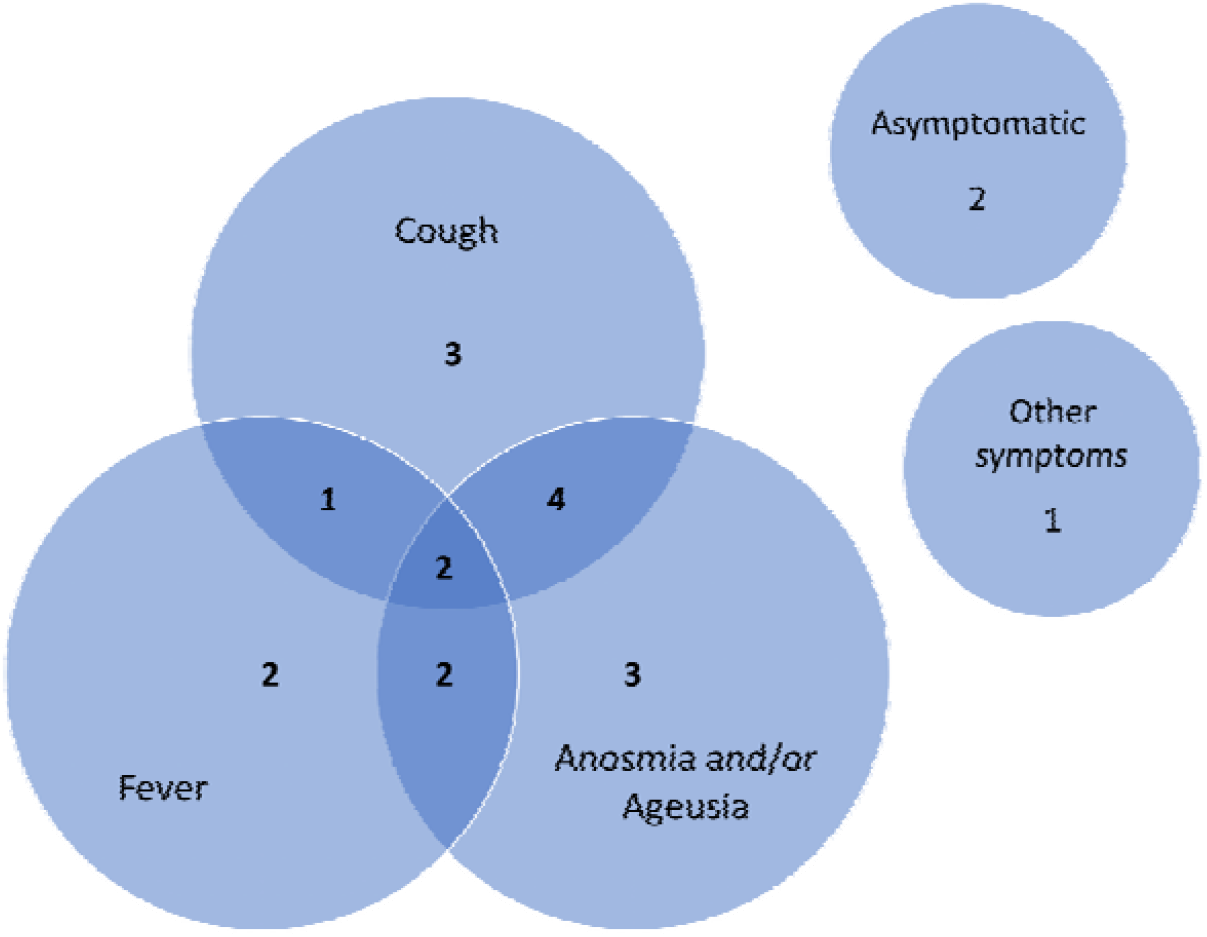
Venn diagram of symptoms in 20 false positive patients in the study by Fournier et al (2020)^11^. This figure was compiled based on the symptoms listed in the manuscript, showing how hallmark symptoms of Covid-19 were being exhibited by patients who had otherwise tested negative with a central gold standard PCR assay. The “other symptoms” patient exhibited eosinopenia, which was also found in 14 of the others, but would not be overtly obvious to patients without a clinical assay.

With false negatives, then here there are a wider range of studies that fall outside the control limits. False positives may be considered an inconvenience for the patient in terms of them and their close contacts being subjected to restrictions on social contact and may also result in delays to treatment if e.g., the patient is being monitored prior to surgery. False negatives, on the other hand, could have much more serious consequences, giving false reassurance to a subject who then may go on to infect countless others. It is therefore important to consider what may be behind those rates shown in Figure 5, and for that we consider the Visseaux et al^19^ and Lubke et al^15^ studies in more detail.

Beginning with Lubke et al., then the high number of false negatives can be easily explained when examining the Cq data helpfully provided by the authors. Of the 15 false negatives, the Cq value of the comparator assay is >35 in 10 of those cases, and so would be beyond the cut-off of assays such as the Roche cobas SARS-Cov-2 test. If these were therefore reassigned as true negatives, then the PPA changes from 83.15% to 93.67%, and the NPA changes from 0% to 90.91%. Furthermore, Lubke have used primers that detect the E gene in a region that overlap with the Corman primers, which were one of the first to be published, based on the limited number of sequences available at the time^3^. The Lubke primers, however, can be seen to have improved detection of target, with a mean Cq decrease of 1, which could feasibly bring some samples of the lowest viral titre to within the detection range of the assay. It is of note that the two studies from Bordi et al., which have a high false positive rate^4, 5^ use the Corman primers as the comparator to the DiaSorin direct-to-PCR assay, and it is therefore feasible that the primers in the DiaSorin assay exhibit a similar capacity to amplify the RNA to within a detectable range.

For the Visseaux et al study^19^, the authors examine three direct-to-PCR methods, with the commercial Sansure assay exhibiting number of discordant well within the control limits. However, the primers used for the Prime direct and Prime script cohorts (both reagents from Takara Bio) are the same primers from the Corman manuscript^3^ which Lubke et al have shown can be improved upon^15^.

Taken together, the reporting of discordant results does need careful consideration, and in the majority of the studies in this meta-analysis, no third resolver assay was used to help determine the correct call for the discordants. It is wise to consider what such a resolver assay should consist of when examining a direct-to-PCR method compared to an extraction-based RT-PCR method. The former may dilute the sample, depending on how the swab is handled (such as the swab being added directly to a lysis buffer, and a small amount taken for PCR), whereas an extraction may cause the virus present in e.g., 200 µl of viral transport medium to be eluted in 50 µl prior to the PCR, therefore enriching it. Making direct comparisons is therefore difficult, but an approach which incorporates a standard curve in the resolver assay is the most sensible approach. This standard curve should include several dilutions of RNA of known copy number which go down to and beyond the limit of detection (LoD), as determined during validation of the resolver assay. This will help establish a Cq cut off for the resolver assay beyond which any discordant samples would be definitively called as negative, with all those with a Cq above the LoD, definitely positive.

The forest plot of the sensitivity and specificity of the cohorts are shown in Figure 7.

**Figure 7.**
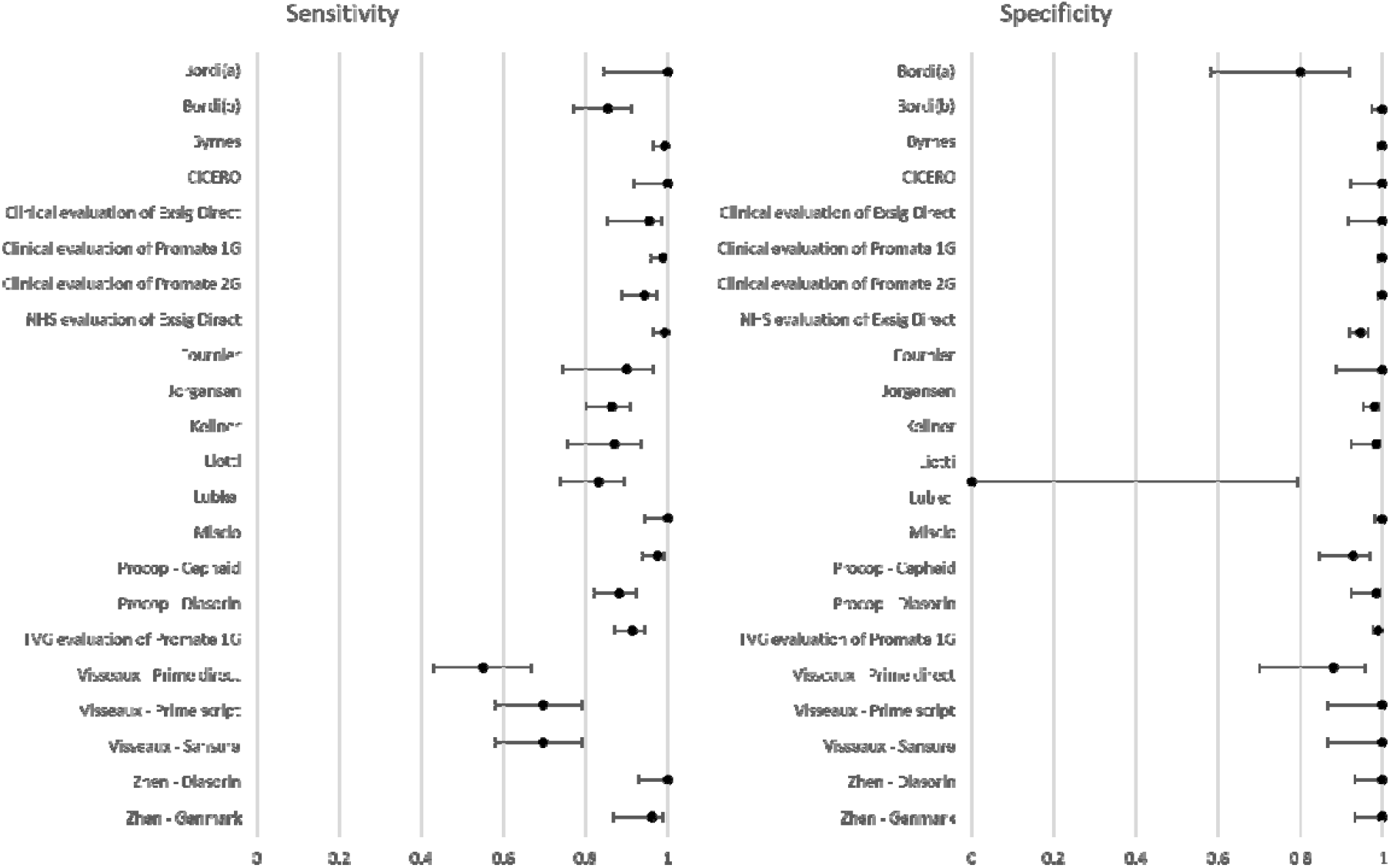
Forest plots of sensitivity and specificity. The dot indicates sensitivity/specificity, with the error bars showing the 95% confidence interval (using the Wilson score interval^21^ due to cohort size and results tending to 0 and 1)

The results from the meta-analysis can also be subgrouped by assay (Table 5)

**Table 5.** Summary of results by assay.

| Assay | Samples | True Positive | False Positive | False Negative | True Negative | PPA | NPA | OPA |
| --- | --- | --- | --- | --- | --- | --- | --- | --- |
| Cepheid<sup>17</sup> | 238 | 163 | 5 | 4 | 66 | 97.61% | 92.96% | 96.22% |
| DiaSorin<sup>4, 5, 14, 17, 20</sup> | 787 | 366 | 14 | 27 | 380 | 93.13% | 96.45% | 94.79% |
| Exsig Direct<sup>7, 8, 10</sup> | 1019 | 328 | 0 | 9 | 682 | 97.33% | 100% | 99.12% |
| Genmark<sup>20</sup> | 104 | 49 | 0 | 2 | 53 | 96.08% | 100% | 98.08% |
| Hyris<sup>16</sup> | 266 | 63 | 0 | 0 | 203 | 100% | 100% | 100% |
| PROmate 1G<sup>8, 18</sup> | 847 | 264 | 5 | 23 | 555 | 91.99% | 99.11% | 96.69% |
| PROmate 2G<sup>9</sup> | 465 | 181 | 0 | 2 | 282 | 98.91% | 100.00% | 99.57% |
| Sansure<sup>19</sup> | 94 | 48 | 0 | 21 | 25 | 69.57% | 100% | 77.66% |
| VitaPCR<sup>11</sup> | 534 | 155 | 20 | 1 | 358 | 99.36% | 94.71% | 96.07% |
| Totals | 4354 | 1617 | 44 | 89 | 2604 | 94.78% | 98.34% | 96.95% |

## Discussion

### Summary of main findings

Rapid direct-to-PCR systems further aim to reduce the speed of testing by combining rapid-PCR instruments with direct-to-PCR assays, to generate results in less than 2 hours. If sufficiently accurate, rapid tests returned within the same clinical encounter can facilitate timely decisions concerning the need for isolation and contract tracing activities^1^ and create novel patient care pathways for the rapid diagnosis and treatment of COVID-19, and support infection prevention and control.

This appears to be the first meta-analysis assessing the accuracy of rapid direct-to-PCR in the detection of SARS-CoV-2 and presents our current state of knowledge in this rapidly expanding field. We intend that our first iteration of this living review will be updated in 3-6 months, as further studies and release of other direct to PCR assays expand the data available.

Through this analysis, several potential sources of heterogeneity in the comparative evaluation methodology, whereby a new RT-PCR assay is compared with a reference RT-PCR assay have become apparent. Firstly, the direct-to-PCR reaction involves a dilution step whereas the more traditional RT-PCR involves a concentration step. In the dilution step, the quantity of nucleic acid in the sample is diluted, potentially increasing the likelihood of a false negative result. Conversely for the same viral load in an extracted PCR reaction, the concentration step concentrates or enriches the quantity of RNA potentially. Therefore, this could present a significant source of systematic bias. Furthermore, comparing assays with different gene targets adds another fundamental source of bias because detection of sub-genomic RNA may not be a suitable indicator of active replication/infection. Sub-genomic RNA was found to have accumulated and remain detectable 17 days after infection.^22^ Therefore, the comparison of an assay that detects genomic RNA (ORF1ab) versus an assay detecting sub-genomic RNA (E-gene) may introduce a further systematic bias.

Nevertheless, our findings show that direct-to-PCR assays perform equivalently to the gold standard centralised laboratory PCR systems for the diagnosis of SARS-CoV-2.

## Data Availability

Data can be made available on request

## Author’s conclusions

This meta-analysis of the accuracy of rapid direct PCR for the diagnosis of SARS-CoV-2 included sixteen reports encompassing 22 datasets with a total of 5322 patient samples (2220 positive). As some of the studies using assays developed in research laboratories were a source of a number of results which biased the overall findings, we make the following conclusions solely on the commercial kits, which encompasses 16 datasets, with a total of 4354 patient samples (1706 positive). Therefore, the overall agreement between the commercial rapid direct PCR and gold standard centralised laboratory PCR was 96.95% with 94.78% positive percent agreement and 98.33% negative percent agreement. The Cohen’s kappa statistical coefficient k = 0.96, and Youden Index = 0.931 indicate almost perfect agreement. These results show that commercial direct-to-PCR assays perform equivalently to the standard centralised PCR systems for the diagnosis of SARS-CoV-2 and support the investigation of new rapid patient pathways for the detection of COVID-19.

## Declarations of Interest

The authors Stephen P. Kidd*, and J.E Martin* have no direct conflict of interest, JEM is principal investigator on the CICERO trial, funded by Novacyt. R.A. Trevor, S. Grippon, L. Koh, H. Chen, U. Jan, D. Borley, M.J Coldwell and P. Oladimeji are employees of Novacyt group.

*joint senior authors

## Appendices

**Appendix 1.** outlines the Direct-to-PCR assays that were analysed in this review

| Study | Direct to PCR assay | Gene target(s) | Comparator assay | Comparator Gene Targets |
| --- | --- | --- | --- | --- |
| **Bordi et al. 2020a** | Simplexa™ COVID-19 Direct (DiaSorin) | ORF1ab and S | Extraction followed by Corman's protocol | E and RdRp targets from Corman's protocol (not N) |
| **Bordi et al. 2020 b** | Simplexa™ COVID-19 Direct (DiaSorin) | ORF1ab and S | Extraction followed by Corman's protocol | RdRP, E and N gene |
| **Byrnes et al. 2021** | Research Assay | N1 and N2 | In house extraction | N1 and N2 |
| **CICERO** | exsig® COVID-19 Direct | ORF1ab | ROCHE cobas® SARS-CoV-2 Test | ORF1ab and E |
| **Clinical evaluation of exsig** | exsig® COVID-19 Direct | ORF1ab | Genesig COVID-19 1G | ORF1ab |
| **Clinical evaluation of PROmate 1G** | PROmate® COVID-19 1G | ORF1ab | Genesig COVID-19 1G | ORF1ab |
| **Clinical evaluation of PROmate 2G** | PROmate® COVID-19 2G | ORF1ab and M | Thermo Fisher TaqPath and Cepheid | ORF1ab, N and S |
| **Exsig NHS validation** | exsig® COVID-19 Direct | ORF1ab | Comparator assay only provided by 1 study centre (Cepheid Gene Xpert SARS CoV-2 Kit) while the other centres did not disclose comparator assays | Cepheid: E and N2 gene: Others did not specify |
| **Fournier et al 2020** | VitaPCR™ SARS-CoV-2 Assay | N (one site unique to SARS-CoV-2, another in the N gene common to SARS-CoV2, SARS-CoV and the SARS-like coronavirus from bat | 2019-nCoV Real-Time Multiplex RT-PCR Kit (Liferiver and Shanghai ZJ Bio-Tech Co., Ltd), | ORF1ab, E and N |
| **Jørgensen et al. 2021** | Research Assay | E (Corman primers/probe) | MagNA Pure 96 DNA and Viral NA small volume kit | E (Corman primers/probe) |
| **Kellner et al. 2021** | Research Assay | N1 | In house extraction | E (Corman primers/probe) |
| **Liotti et al. 2021** | Simplexa™ COVID-19 Direct (DiaSorin) | ORF1ab and S | Allplex™ 2019-nCoV | RdRP, E and N gene |
| **Lübke et al. 2020** | Research Assay | E (overlap with Corman E) | RNA-based RT-qPCR-protocol by Corman OR by | ROCHE cobas® SARS- |
|  |  |  | the cobas® SARS-CoV-2 test on the cobas® 6800 system (Roche) | CoV-2 Test targets ORF1ab and E gene |
| **Miscio et al. 2020** | bKIT Virus Finder COVID-19 (Hyris) | N | Abbott Kit: Real-Time SARS-CoV-2 and Roche Kit: LightMix®Modular SARS-Cov-2 (COVID19) | ROCHE cobas® SARS-CoV-2 Test targets ORF1ab and E gene |
| **Procop 2021 - Cepheid** | Xpert Xpress SARS-CoV-2 (Cepheid) | E and N2 | CDC 2019 nCoV Real-Time RT-PCR Diagnostic Panel | N1 and N2 |
| **Procop 2021 - Diasorin** | Simplexa™ COVID-19 Direct (DiaSorin) | ORF1ab and S | CDC 2019 nCoV Real-Time RT-PCR Diagnostic Panel | N1 and N2 |
| **PROmate TVG validation** | PROmate® COVID-19 | ORF1ab | Hologic Panther SARS-CoV-2 assay | ORF1ab |
| **Visseaux 2021 - Prime direct** | Research Assay | E and RdRp (E are the Corman primer/probe) | RealStar® SARS-CoV-2 RT-PCR Kit 1.0 and the Cobas® SARS-CoV-2 kit used on the Cobas® 6800 system. | E and S |
| **Visseaux 2021 - Prime script** | Research Assay | E and RdRp (E are the Corman primer/probe) | RealStar® SARS-CoV-2 RT-PCR Kit 1.0 and the Cobas® SARS-CoV-2 kit used on the Cobas® 6800 system. | E and S |
| **Visseaux 2021 - Sansure** | SARS-CoV-2 SANSURE®BIOTECH Novel Coronavirus (Sansure) | N and ORF1ab | RealStar® SARS-CoV-2 RT-PCR Kit 1.0 and the Cobas® SARS-CoV-2 kit used on the Cobas® 6800 system. | E and S |
| **Zhen et al. 2020 - DiaSorin** | Simplexa™ COVID-19 Direct (DiaSorin) | ORF1ab and S | CDC, DiaSorin and Hologic - positive in any one is considered overall positive | Multiple |
| **Zhen et al. 2020 - GenMark** | ePlex SARS-CoV-2 test (GenMark) | N | CDC, DiaSorin and Hologic - positive in any one is considered overall positive | Multiple |

